# Collective Health Decision-Making: Exploring Examples from Chinese Families in Australia

**DOI:** 10.1101/2025.10.14.25338024

**Authors:** Bingyi Han, Shanton Chang, Dana McKay, Carlene Wilson, Joyce Jiang, Jennifer McIntosh, Mark Jenkins

## Abstract

Australia’s patient-centred healthcare system emphasises individual autonomy, but this approach may not align with families that have culturally collectivist values. One example is Chinese culture, where collectivist values are rooted in Confucian principles of responsibility. Chinese people are the largest non-English-speaking migrant group in Australia, so understanding how Chinese families make health decisions is important for improving communication, access, and participation in preventive care. This study examined how Chinese immigrant families in Australia make health-related decisions, focusing on vaccination and bowel cancer screening, and how cultural and intergenerational factors shape these processes. We conducted semi-structured interviews with 13 Chinese multigenerational families living in Victoria in 2023, each including at least two generations living under one roof. Interviews were conducted in Mandarin for each family with one older parent, one adult child, and both together, and were analysed using reflexive thematic analysis. Four themes were identified: (1) language and cultural alignment, with families relying on Chinese-speaking doctors and culturally familiar information sources; (2) collective effort and interdependence, with adult children often leading or supporting decisions due to English proficiency and familiarity with the healthcare system; (3) intergenerational conflict, particularly when older parents withheld health concerns to avoid burdening their children; and (4) negotiation and implementation strategies by adult children, including using doctors’ authority and arranging appointments. Decision-making patterns ranged from full delegation (to adult child) to shared deliberation, with respect for parental autonomy remaining important. These findings highlight the interplay between cultural norms, family roles, and systemic barriers to accessing healthcare. We demonstrate that strict adherence to Western privacy norms limits access to care for collectivist families. Instead, healthcare services should provide accessible language support, culturally tailored health education, and options for both family-based and independent patient engagement to improve health outcomes in culturally and linguistically diverse populations.

## 1. Introduction

The Australian health system emphasises a patient-centred approach [1], a paradigm that underscores the importance healthcare institutions place on an individual’s right to choose and their need to participate in decisions made about their care [2]. This approach seeks to ensure that individuals accessing health care services are treated with respect and responsiveness [3]. It also emphasises the importance of individuals being provided with information and resources about relevant matters, specifically, risks and benefits, so they can make informed decisions about their health. Achieving shared decision-making requires that individuals are equipped with the necessary medical information, knowledge of risks and benefits, and are actively supported throughout the decision-making processes to ensure decisions truly reflect their values and preferences [1, 4].

Research and anecdotal evidence suggest that the patient-centred approach, which advocates individual autonomy in decision-making, may not fully align with patients’ decision-making preferences in some cultural contexts. Indigenous Australians, for instance, frequently approach health care through a lens of community well-being, where decision-making is deeply rooted in the interconnection of the family and the wider community [5]. Similarly, within Chinese households, there is a prevalent expectation for family members to be deeply involved in others’ healthcare decisions [6, 7]. These patterns of collective decision-making, common among families from collectivist culture, represent the health decision making practices of a significant demographic in Australia that is very different from the preference for individualistic decision-making that is otherwise widespread in Western communities.

The paradigm of collective decision-making acknowledges the collective group (families, communities, or other groups) as a central cohesive unit [8]. Far from being a mere aggregation of individual preferences, collective decision-making is deeply rooted in the ethos of interdependence, shared responsibility, and mutual support [9–11]. Collective decisions are influenced by the group’s shared values, knowledge, and experiences, and are aimed at optimising the health outcome of the group and its members [8]. In such settings, the well-being of an individual is linked to the health of the family group and collective harmony [12, 13].

Chinese families, who are typically collectivist in their cultural outlook, are the largest non-English speaking migrant group in Australia; making up 5.5% of the total population [14]. Australia’s individualistic [12] healthcare ethos and Chinese cultural values may represent contrasting paradigms, creating complex dynamics for Chinese families. Within Chinese families, medical decisions are often made collectively [9, 15], with members acting as intermediaries between the patient and doctors, assisting in treatment planning, sharing medical costs and bearing medical consequences [10, 16]. Migrating to Australia exposes Chinese families to Western individualist norms, which may broaden perspectives but also create tensions when health concepts and expectations diverge [17–19]. Intergenerational differences can further complicate decision-making; parents may expect children to uphold heritage cultural practices while younger generations adopt Western norms [17, 20]. These differences influence how health information is shared and how collective decisions are negotiated, and challenges family communication and practices [20].

Although previous research highlight cultural adaption identify negotiation and family involvement in healthcare [17, 20, 21], little is known about how Chinese families in Australia undertake health care decision-making, especially when individual responsibility needs to be balanced against communal obligation. Understanding these dynamics is critical because Mandarin-speaking Chinese communities in Australia have historically shown lower participation than the national average in preventive health programs (e.g., [22]). This research therefore asks: *How do Chinese families in Australia, living in extended family households, interact, negotiate and arrive at healthcare decisions?*

## 2. Methods

We conducted in-depth interviews in 2023. We recruited 13 Chinese families in the state of Victoria, Australia, each spanning at least two generations living under the same roof. The interviews were with one adult child (aged 18 years or above, born in China), one aging parent (aged 50 years or above, born in China), and a joint group of family members to gain insights into their decision-making processes. We explored their attitudes and approaches in two health decisions: COVID 19 vaccination and bowel cancer screening. Vaccination serves as an example to explore family decision-making patterns as it is a recent and relatable health decision that most families have encountered [23]. Bowel cancer screening was included due to the notably lower participation rates among people who speak a language other than English at home, highlighting the need to understand the cultural factors influencing this decision [22]. This research was approved by the Human Research Ethics Committee of the University of Melbourne (Ethics ID number: 26567).

### 3.1 Data Collection

In this section, we address our data collection approach, including participant recruitment.

#### Participant Recruitment

Adult children aged 18 years or above, and their parents, aged 50 years or above were eligible for inclusion. All participants were originally from China and lived as a member of a multi-generational family in Victoria, Australia. Participants needed to be able to communicate effectively in either English or Mandarin.

Initially, in August 2023, potential participants were identified from within the research team’s personal networks within the Chinese community. Three methods were used to approach potential participants: (i) individuals were contacted via email to gauge their interest in participating and were invited to register via a secure online survey, (ii) we leveraged our professional networks to disseminate the online registration forms, and (iii). digital flyers were created featuring a QR code linked to the online registration form and were shared via local WeChat group chats, and printed flyers were placed in Chinese restaurants and supermarkets across various locales in Melbourne.

Sixty-seven expressions of interest, received online, were followed up through email or phone calls to verify eligibility based on our inclusion criteria. Of these, 32 individuals’ families met the criteria. To ensure diverse data sources, we finalized a cohort of families from various suburbs in Melbourne. This sample size was deemed adequate based on prior research recommendations [24, 25]. With two exceptions (Family 6 and Family 10, where only an older parent and an adult child lived together), our participant families comprised 3 generations (grandparents, their adult children, and grandchildren). Table 1 presents the participant information.

**Table 1.** Participants Information. – * In Family 6 and Family 10, two generations live together without grandchildren. 26 people participated the interviews-two from each family, each from a different generation.

| Family | Living situation | Interviewees | Mode of interview | Post code of interviewees |
| --- | --- | --- | --- | --- |
| 1 | 2 grandparents,<br>2 adult children,<br>2 grandchildren | 1 grandparent,<br>1 parent | In person | Vermont VIC 3133 |
| 2 | 1 grandparent,<br>2 adult children,<br>1 grandchild | 1 grandparent,<br>1 parent | In person | Vermont VIC 3133 |
| 3 | 2 grandparents,<br>2 adult children,<br>2 grandchildren | 1 grandparent,<br>1 parent | In person | Templestowe VIC 3106 |
| 4 | 2 grandparents,<br>2 adult children,<br>1 grandchild | 1 grandparent,<br>1 parent | Zoom | Glen Waverley VIC 3150 |
| 5 | 2 grandparents,<br>2 adult children,<br>3 grandchildren | 1 grandparent,<br>1 parent | In person | Doncaster VIC 3108 |
| 6 | 1 older parent,<br>1 adult child* | 1 older parent,<br>1 adult child* | Zoom | Box Hill VIC 3128 |
| 7 | 2 grandparents,<br>2 adult children,<br>2 grandchildren | 1 grandparent,<br>1 parent | Zoom | Glen Waverley VIC 3150 |
| 8 | 2 grandparents,<br>2 adult children,<br>1 grandchild | 1 grandparent,<br>1 parent | Zoom | Wantirna South VIC<br>3152 |
| 9 | 2 grandparents,<br>2 adult children,<br>1 grandchild | 1 grandparent,<br>1 parent | Zoom | Mt Waverley VIC 3149 |
| 10 | 1 older parent,<br>1 adult child* | 1 older parent,<br>1 adult child* | Zoom | Templestowe VIC 3106 |
| 11 | 1 grandparent,<br>2 adult children,<br>2 grandchildren | 1 grandparent,<br>1 parent | Zoom | Box Hill VIC 3128 |
| 12 | 2 grandparents,<br>2 adult children,<br>2 grandchildren | 1 grandparent,<br>1 parent | Zoom | Ringwood VIC 3134 |
| 13 | 1 grandparent,<br>2 adult children,<br>2 grandchildren | 1 grandparent,<br>1 parent | Zoom | Croydon, 3136 |

#### Procedure

Potential participants were provided with a plain language statement and a consent form. Upon receipt of their signed consent forms, we invited participants to complete a pre-interview questionnaire. This preliminary survey gathered basic demographic information, including age and family composition, such as the number of generations cohabitating. To express our gratitude for their contribution, each participating family was presented with a $100 supermarket gift card.

Interviews were conducted either by in-person visits by the primary researcher to the participant’s residence or through virtual meetings via Zoom, according to participants’ preferences. For each family, we first interviewed one grandparent alone for 30 minutes then interviewed one of their adult children for 30 minutes. This allowed the grandparents a rest period before participating in the subsequent joint session. When there was more than one eligible grandparent or adult child, the family themselves nominated the person who would participate on behalf of that generation. We conducted a joint interview involving one adult child and one grandparent for approximately 30 minutes. All interviews were conducted in Mandarin, according to participant preference.

At the beginning of each interview, we briefed participants on the research focus and obtained informed consent for deidentified data collection. We then engaged participants in discussions about their family’s health decision-making processes, specifically exploring their experiences during the pandemic with vaccination decisions, and bowel cancer screening practices. It is important to note that our research does not advocate for or against any particular health choices, such as vaccinations or cancer screening. Our objective was to understand the dynamics of decision-making within families without bias or judgment.

The interview period spanned August 20th to October 10^th^ 2023. Session durations ranged from 46 minutes to 1 hour and 38 minutes, averaging 57 minutes per family (see Table 1). All interviews were audio-recorded and transcribed by the first author before the analysis. All data were analysed in anonymised form. The authors had no access to any information that could identify individual participants after data collection. All identifying details were removed or coded prior to analysis to ensure participant confidentiality.

### 3.2 Data Analysis

Utilising reflexive thematic analysis, our approach followed the 6-phases suggested by Braun and Clarke [26]. Analysis was led by the first author, who is fluent in Mandarin and familiar with Chinese and Australian culture.

In phase 1, the first author conducted an initial review of the interview transcripts. This phase involved distilling the transcripts into a more concise format, generating initial ideas of the family discussion patterns and describing information-seeking and sharing behaviours. In phase 2, the first author coded the condensed interview transcripts at the sentence level using Microsoft Word. In phase 3, the research team arranged the initial codes into potential themes, with their interrelations sketched through mind maps [26]. In phase 4, the research team discussed the mind maps and reflected and refined the theme structure. In phase 5, the themes were further defined and categorised, sub-themes were consolidated. In phase 6, we drafted the initial report, which further revised the themes. Following this, the research team revisited phases 3 to 6 in an iterative cycle, refining the themes. This iterative process is in line with the principles of reflexive thematic analysis [27], ensuring a progressively recursive analysis and interpretations [28, 29].

## 3. Findings

In this section, we present results, which highlight the complex interplay of cultural, generational and systemic factors that influence health decision-making within Chinese families living in Australia. We generated five primary themes from our analysis, as presented in Table 2 below. The findings within each theme are summarised in the following sub-sections. While we acknowledge that quantifying codes is atypical in reflexive thematic analysis, we include counts here to enhance transparency and illustrate the relative salience of patterns across our sample. Given the exploratory nature of our study and the modest sample size of 13 families, our goal is not to generalise but to foreground recurring dynamics and shared strategies that emerged across diverse family contexts.

**Table 2.**
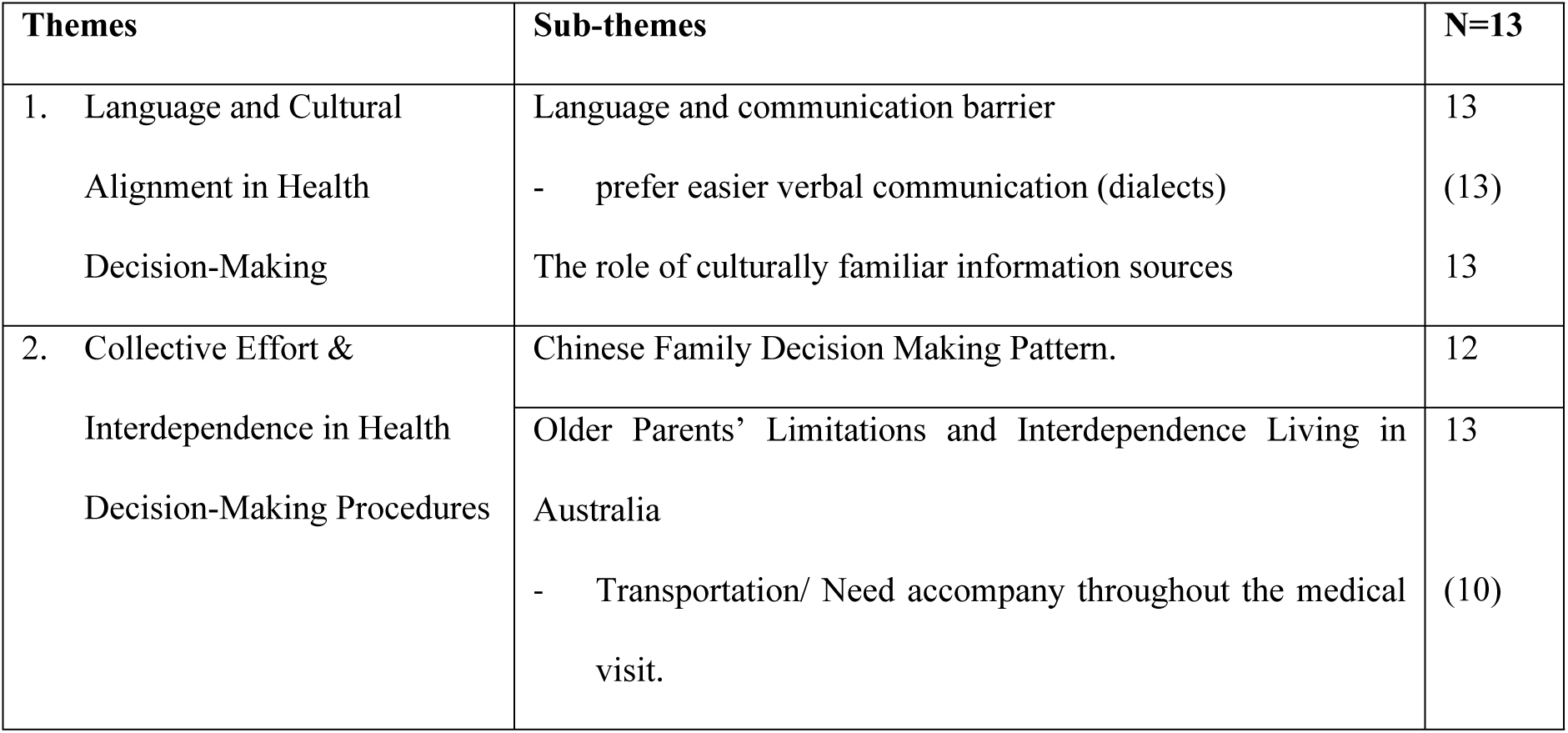
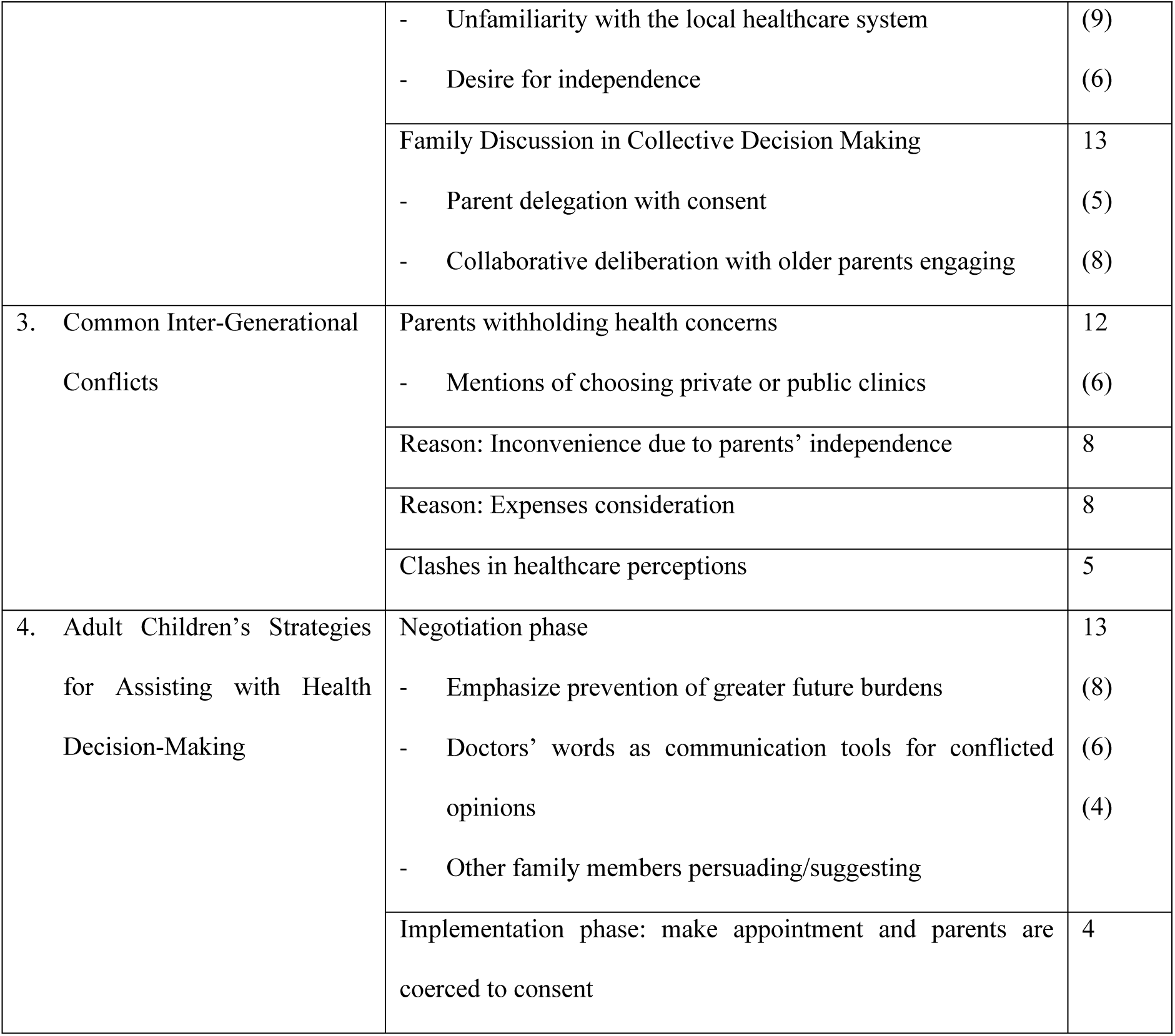
Identified Themes. – * N = number of families in which the theme was identified. A theme was counted if it appeared in either the parent, the adult child, or the joint family interview. Parent and child perspectives were not required to align; differing views within the same family were still coded under that theme if both related to the topic.

### 3.1 Language and Cultural Alignment in Health Decision-Making

This theme explores how language barriers and the need for culturally aligned information sources impact health-related decision-making among Chinese families in Australia. Access to information and healthcare providers who speak their language or share cultural backgrounds enhances the ability of these families to make informed health decisions. This section combines insights into the significance of language accessibility and the reliance on culturally familiar sources of information.

#### 4.1.1 Language Barriers and Access to Health Professionals

Families reported that language barriers were a significant obstacle when accessing healthcare and understanding medical information. Although this challenge was most pronounced among older family members with limited English proficiency, adult children also reported difficulties, especially with technical medical terminology.

Consequently, many families prioritised finding Chinese-speaking General Practitioners (GPs) or specialists, with 4 families even seeking doctors who shared their regional dialect to facilitate clearer communication.

One participant expressed the need to access information in Chinese as a priority:

> “Actually, I also encounter language limitations, despite my fluency in the workplace. Terms related to medical conditions are sometimes unfamiliar to me, I need to translate or do additional research for full comprehension. It takes effort. And this effort might only be reserved for instances where I’m keen to understand, for example, what new policies the Australian government is promoting on English websites, or discussions from English-speaking online communities. Otherwise, I just prefer Chinese information sources.” (F7 adult child)

The preference for Chinese-speaking healthcare providers was also motivated by the desire for direct, transparent communication, which was particularly important for older family members who valued autonomy in understanding their health:

> “Since my mother doesn’t speak English and isn’t fluent in Mandarin, we mostly choose Cantonese-speaking family doctors, for ease of communication fitting her language preferences. She is willing to communicate with doctors, and wants to have a transparent, clearer, and more direct understanding of her own condition… As I’m often unavailable to accompany her to visit doctors due to full-time work commitments, a Chinese-speaking doctor or even a Cantonese-speaking doctor can enable her to manage appointments independently.” (F6 adult child)

#### 4.1.2 The Role of Culturally Familiar Information Sources

Families frequently relied on information from friends and Chinese-language social media platforms. This preference was not only a matter of convenience but also reflected a need for cultural familiarity and relatability.

Among most of the families interviewed, both adult children and older parents identified friends’ recommendations and shared experiences as their primary source for health information. Most families indicated that their social circle uses the same GP, attends the same Chinese clinic, and opts for the same insurance type and provider. The decision-making process was heavily influenced by word-of-mouth recommendations within their social network.

This significant influence from friends was prominent during critical health challenges, such as those presented by the pandemic:

> “My parent’s opinions and decisions about vaccinations were significantly influenced by their friends’ choices and the brands they opted for… Those decisions are often made after our discussion with our friends…” (F9 adult children)

Many families, especially older parents, trusted information sourced from Chinese media platforms such as WeChat (articles from WeChat Official Account, posts on WeChat Moments, contents shared in WeChat group chat), search results from Baidu and Xiaohongshu, and other Chinese media, because these sources were more accessible and aligned with their cultural context. Among the various Chinese-language information sources, user-generated educational and explanatory content was popular and was often referred to by the interviewed families.

> “Despite the availability of officially sanctioned health news and information from accredited Australian sources in translated Chinese, the language used is often excessively formal and not aligned with everyday speech. Individuals typically search using informal, colloquial language or specific keywords, leading to these official resources being ranked lower by search engines and thus, hard to locate. Moreover, the formal nature of the language can make comprehension challenging. So we tend to favour more approachable, layman-friendly educational and explanatory content created by other Chinese online users.” (F7 adult child)

As described by F7 adult child, the overly formal language of official resources not only hindered comprehension but also created a disconnect between the information provided and the way families naturally seek health guidance. This misalignment drove them to prefer user-generated content, which was perceived as more relatable and easier to understand. Nevertheless, adult children expressed concerns about the credibility of some of these sources, especially for older parents who might not scrutinise information rigorously:

> “Exaggerated headlines on WeChat are designed to capture attention and gain revenue. The scattered information, often not from verified medical sources, can be misleading or completely false. But older individuals tend to trust what they find online without scrutiny.” (F12 adult child)

A particularly telling incident occurred during a family interview where an older parent asserted his independence in sourcing information, trusting online professional advice over that of his daughter’s, leading to a palpable disagreement:

> “…by professional information, he (my father) means the article he browsed on WeChat, Baidu, or even from the commercial advertisement - Like those claiming bitter melon tea lowers blood sugar on the product website. He trusts any information displayed online. He also cited his friends’ usage as validation, despite the dubious origins of such information.” (F5 adult child)

Although adult children often played a role in validating information for their parents, participants also noted the importance of formal community events and resources provided in Chinese by local councils and healthcare institutions. Such initiatives were seen as valuable for providing credible information and for fostering social connections and mitigating isolation:

> “This could bridge the gap in their fragmented and sometimes questionable online information sources, empowering them to make independent decisions without overly relying on their children…It not only represents a learning journey for the elders, it serves as an avenue to fill knowledge gaps with credible information, and it provides more than just education, but creates community engagement opportunities for social connectivity, which is crucial for the mental health of elders in an unfamiliar foreign social environment. (F5 adult child)

### 3.2 Collective Effort and Interdependence in Health Decision-Making Procedures

This section describes the participants’ perceptions of decision-making in Chinese families, showing a clear tendency towards a collective approach.

#### 4.2.1 Chinese Family Decision-Making Patterns

In Chinese families, health decision-making often followed a collective process, as summarised in Figure 1.

**Fig. 1.**
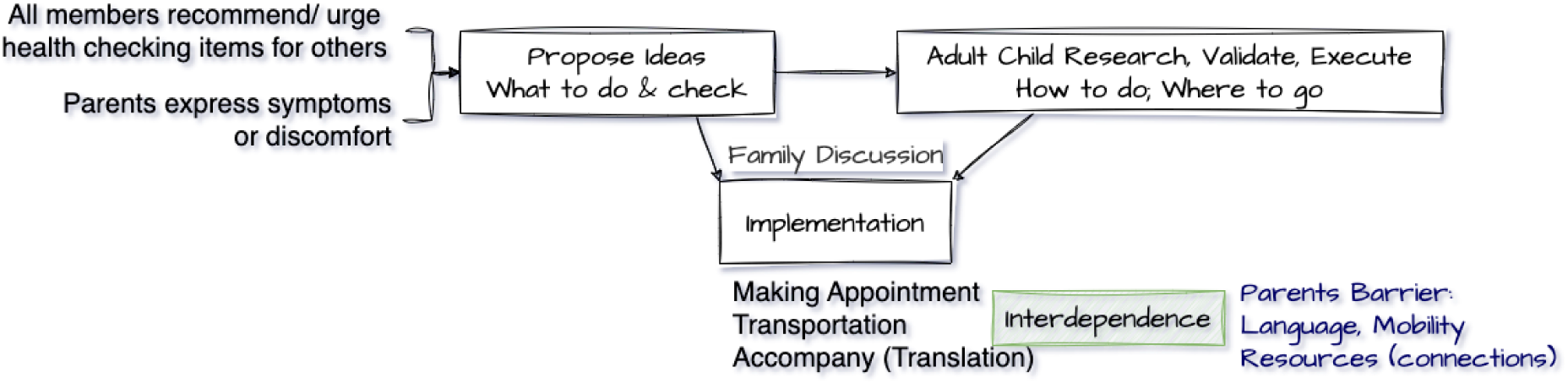
Chinese Family Decision-Making Process. The figure depicts family health decision-making stages sequentially, but in practice these steps often overlapped. In particular, the early phases of proposing ideas and validating information were usually embedded within ongoing family discussions, rather than occurring as distinct steps outside of the discussion.

The decision-making process was usually initiated when a family member, usually a parent, voiced their health discomforts or symptoms, and proposed actions to address it. Alternatively, there was also mutual prompting within the family to maintain regular health check-ups. For instance:

> “When I feel unwell, I inform my daughter, who then advises me and accompanies me to the doctor… I also reminded my daughter of her health check-ups, like during her pregnancy when she was a focus of the family, I inquired about her upcoming routine visit. In our family, the individual in the health situation requiring care becomes the focal point of attention and support from the rest. (“谁生病, 谁就是被关注的主体”)” (F7 parent)

Following the initial proposal for a health-related action, the adult child typically assumed responsibility for gathering information and organising care. They relied on consultations with GPs or friends to validate decisions, and their role extended to arranging appointments and accompanying their parents, mainly to assist with language translation. In most families, parents deferred to their children due to the latter’s familiarity with the Australian healthcare system, although older family members like F1’s father, may still contribute authoritative health knowledge due to prior experience. Here is an example from the educated father and the child assisting with implementation:

> “I suggest important health checks for each family member. My daughter then locates credible facilities for these checks, arranges the appointments, and takes us there. It is mainly the language limitations that I couldn’t do a lot of things by my own” (F1 parent)

> “My father has worked in the health sector before retirement in China. We seek health advice from him as we rely on his expertise; So I take these suggestions and put them into action.” (F1 adult child)

#### 4.2.2 Older Parents’ Limitations and Family Dependence

Older parents in Chinese families faced distinct challenges when navigating the Australian healthcare system, which often led to a reliance on their adult children. While language barriers were a well-established obstacle (as discussed in Section 4.1.1), they further limited older parents’ actions and independence. Ten families reported that their adult children not only drove them to clinics but also accompanied them. This arrangement was less about transportation challenges and more about overcoming language barriers that made independent travel and healthcare access difficult, requiring children’s assistance:

> “I spend most of my time with my daughter in Australia. Because I don’t speak English, I can’t go many places without her… I rely on her for many things… I can manage on my own only in situations where no English is needed, like having an acupuncture appointment booked at a Chinese medicine clinic.” (F11 parent)

The complexity of the Australian healthcare system, especially in contrast to the systems they were familiar with in China, placed older parents at a disadvantage position. Many struggled with tasks such as managing insurance, booking appointments, and procuring medication, which can be daunting without local knowledge.

> “The entire procedure, from GP appointments to surgery, differs from what I knew in China. I let my child make decisions for me regarding insurance and healthcare. I simply follow her lead.” (F3 parent)

In addition, older parents lacked the social connections within the healthcare community that they once had in China, which further limited their independence.

> “I rely on my children because I don’t have a network here. Back home, we always had friends in hospitals. Here, I have to depend on my child for everything, including understanding insurance and finding doctors.” (F4 parent)

Despite these constraints, some older parents expressed a desire for greater autonomy. They highlighted how dependence on their children can be inconvenient for both parties, especially when language barriers or system unfamiliarity prevent them from managing appointments alone.

> “I would prefer to attend appointments independently, but without Chinese language support, I have no choice but to rely on my daughter. I want to reduce her burden, but it’s not feasible.” (F9 parent)

These reflections revealed a tension between parents’ wish for independence and the practical limitations that compel them to rely on family support. Although progress was being made, as some parents gradually familiarised themselves with local procedures, the need for family assistance remained significant.

> “They’re learning the process and can now visit our family doctor independently, which has reduced my involvement somewhat. But for anything complex, they still rely on me.” (F5 adult child)

#### 4.2.3 The Role of Family Discussion in Collective Decision Making

Health-related decisions in Chinese families were often reached through informal discussions during everyday activities, such as cooking or dining. Rather than formal meetings, family members engaged in casual conversations where each member’s input was valued.

> “After seeing the GP, if a decision needs to be made, we discuss it casually while cooking or eating, not in a formal way. Sometimes, we even share videos or articles in our family WeChat group to spark discussion.” (F7 parent)

These discussions were driven by a shared commitment to the health and well-being of all family members, embodying a sense of mutual responsibility. In this context, two predominant patterns of family discussion emerged: **Parental Delegation** and **Collaborative Deliberation**. In the **Parental Delegation** approach from five families, parents often delegated decisions to their adult children. They expressed trust in their children’s judgment and provided consent after suggestions were made, without engaging in extensive deliberation.

> “I trust my daughter completely. She advises me on what to do, and I follow her recommendations. I just inform and consent.” (F12 parent)

> “I take care of [my] parents more often. They trust my judgment, so I handle most health-related tasks.” (F7 adult child)

Conversely, **Collaborative Deliberation,** a theme highlighted by eight families, involved more inclusive discussions, where parents actively participated in information-gathering and shared insights with their adult children. This approach fostered teamwork, with each family member contributing their perspectives and knowledge, leading to a joint decision.

> “Our process is very democratic. Each person brings information to the table, and we discuss it together. Decisions like vaccinations involve everyone, and we reach a consensus without conflict.” (F4 parent)

Regardless of the approach, respect for individual autonomy remained a central value. Even within collaborative discussions, families respected personal preferences and avoided overriding individual choices. In this way, collective decision-making often functioned as a supportive process to inform the individual who retained the authority to decide. For example, although adult children might facilitate parental understanding of health decisions by translating or explaining medical information, final decisions were often left to the person directly affected.

> “Each person makes their own choices. We offer advice, but it’s up to the individual to decide. Our role is to assist, not to direct.” (F7, older parent)

This balanced dynamic allowed for flexibility, enabling the family to adapt to various health situations while ensuring that decisions were made with consent and mutual support.

Within the family discussion dynamics, a notable **role reversal** emerged, where adult children often assumed a leading role in healthcare decision-making, effectively placing them in a guardian-like position. This shift was primarily driven by the adult child’s familiarity with the Australian healthcare system and their proficiency in English, which positioned them as key facilitators. Consequently, they took on responsibilities that range from arranging appointments to overseeing medication management.

> “… I manage my mother’s medications now, and she relies on my decisions here, unlike back in China where she adjusted her doses on her own. I couldn’t control this when she was in China, but now that she’s in Australia, I’m the one buying her medications, she can’t really object to me. (在国内的时候我管不了她, 现在她在澳洲, 药是我在买, 她也反不了我)” (F11, adult child)

Older parents acknowledged this changing power dynamic, sometimes expressing a sense of being “managed” by their children, who now lead family health decisions.

> “Our children make most health decisions themselves now, and it feels like they’re the ones ‘managing’ us.” (F8 parent)

Despite this, parental consent remained crucial, blending traditional respect with the new power dynamics.

### 3.3 Common Intergenerational Conflicts: Not Wanting to Inconvenience Others (添麻烦)

Across families, a recurring source of intergenerational conflict was older parents’ reluctance to voice health concerns, driven by a desire to avoid inconveniencing their children. This reluctance often led parents to silently endure symptoms, delaying medical visits due to concerns over financial costs and the time their children might spend assisting them. Adult children frequently expressed frustration at this reluctance because it sometimes resulted in worsening conditions.

> “My parents tend to endure their discomfort till they can’t bear it (“ 能忍则忍”) and delay treatment, possibly to avoid burdening us with time and effort for doctor visits… My mother delayed seeking treatment for her injured hand, hoping it would recover on its own…” (F11 adult child).

Financial considerations also influenced parents’ hesitance. Although children were willing to cover costs, many parents were wary of the expenses, especially when deciding between public and private healthcare. In some cases, this financial caution may also relate to residency status – some older parents were not eligible for Medicare due to their visa type, which limited access to publicly funded healthcare and may increase out-of-pocket costs even in the public system. Whether to opt for private or public clinics became “a common generational problem or conflicts for Chinese immigrant families” - as noted by family 7 adult child – “… public being cheaper but with longer waits, and private being more expensive but quicker. Elders always suggested that ‘no rush, we can wait’, but we, the younger prefer early intervention for them.”

> “My father doesn’t want to impose a financial burden on me. Even when we insist on early treatment, he worries about the costs with private clinics. I don’t care how much it cost, I just want to cure his illness.” (F4 adult child)

Cultural differences further exacerbated conflicts. Parents often adhered to the Chinese ethos of self-sacrifice for family well-being, enduring ailments to avoid perceived inconvenience. This attitude, rooted in collectivism, clashed with their children’s preference for proactive healthcare.

> “My mother endured a toothache until it was unbearable, not wanting to trouble me for transportation and translation. In Chinese culture, there’s a focus on collective well-being over immediate personal care. (“先人后己,先考虑家庭集体利益”).” (F5 adult child).

In addition, traditional Chinese medical beliefs often conflicted with the Western approaches their children preferred. Older parents expressed scepticism about treatments like physiotherapy and psychological counselling, sometimes viewing them as unnecessary. Differences in the approach to antibiotics and other medications also surfaced. Participants expressed that in China, many antibiotics can be purchased directly from pharmacies without a prescription, whereas in Australia a formal doctor’s consultation and prescription are always required. Parents who were accustomed to this immediate access in China found the Australian process cumbersome and slow..

> “In China, my parents were used to getting antibiotics quickly. Here, the slow process of seeing a doctor frustrates them. They don’t understand the need for appointments and prescriptions to get antibiotics, which can delay care by weeks.” (F12 adult child)

These intergenerational conflicts highlighted the tension between traditional values and the realities of navigating a different healthcare system. Interviewees suggested that some parents needed to balance their cultural inclinations against the logistical and financial constraints of healthcare in Australia.

### 3.4 Adult Children’s Strategies for Assisting with Health Decision-making

In navigating healthcare decisions with older parents, adult children employed various strategies, beginning with negotiation to address their parents’ reluctance and continuing through implementation to ensure follow-through.

#### Negotiation Strategies

Negotiations among family members often involved addressing older parents’ hesitation to disclose health concerns and seek medical care. Children frequently employed persuasive tactics, focusing on the long-term benefits of proactive healthcare to counter parents’ reluctance to “inconvenience” their families. They emphasised that early intervention could prevent more severe and costly health issues down the line.

> “We encourage them not to hide their discomfort to avoid burdening us. Delaying treatment can lead to greater expenses and worse health outcomes. I tell my father that it’s better to address health issues early so it doesn’t become a bigger problem.” (F4, adult child)

Adult children also leveraged an internal cultural logic grounded in collective wellbeing, where an individual’s health is valued, not only for personal benefit, but for its contribution to the family as a whole. They framed early treatment as a way for older parents to maintain their active roles within the household, reinforcing their ongoing importance to family life.

> “We explain that early treatment means faster recovery, so they can continue helping around the house, like picking up the grandchildren. This approach often convinces them of the importance of addressing issues promptly.” (F7, adult child)

When parents expressed fear of medical procedures, or unfavourable diagnoses, children turned to emotional support strategies. In some cases, they involved another trusted family member, often a spouse, to provide reassurance and comfort.

> “My father was afraid of a cystoscopy, so I asked my mom to help persuade him. Her emotional support, along with our assurances, finally convinced him to go through with the procedure.” (F5, adult child)

#### Implementation Strategies

Once consent was obtained, adult children took charge of implementing healthcare plans, sometimes even “coercing” their parents into action by making appointments on their behalf. This practical approach helped overcome initial resistance, as parents often agreed out of respect for their children’s efforts.

> “I made the appointment for my mother and told her she had to go because I had already arranged everything. I even told her I’d paid, so if she didn’t go, we’d lose the money. This compelled her to consent.” (F8, adult child)

In situations where conflicts over medication or treatment arose, adult children often relied on the authority of healthcare professionals to reinforce their recommendations. Parents generally trusted doctors more than their children, which made the doctor’s advice an effective tool for resolving disputes.

> “My father doesn’t take my advice about his diabetes, but he listens to the doctor. Despite my efforts, he only trusts their guidance. It would help if medical authorities could deliver information directly to him, so he wouldn’t need to verify everything.” (F5, adult child)

To provide reliable health information, some adult children suggested that trusted medical organisations should offer information directly through platforms like WeChat, where older parents frequently seek news.

> “If reputable medical institutions could share accurate health information on platforms like WeChat, parents might trust it more. This would spare us the challenge of acting as intermediaries and battling misinformation.” (F5, adult child)

In this structured approach, adult children employed a blend of persuasive negotiation tactics and decisive implementation strategies to support their parents’ healthcare needs, balancing respect for their autonomy with the practical necessities of navigating the healthcare system.

## 4. Discussion

This section discusses collective decision-making, the interaction between the health system and Chinese families, the nuances of families, and provides practice implications designed to support optimal health decision-making among Chinese Australians.

### 4.1 Collective health decision-making among Chinese families in Australia

Our interviews suggest that information-seeking behaviour among older Chinese adults often involve other family members, with the inability to speak the hosting country language a compelling motivator for enlisting others. Notwithstanding reported family involvement, some in the parent generation expressed a strong desire to seek information independently, including engaging with healthcare providers and seeking local information. However, when language limited their ability for direct engagement, children acted as intermediaries. These children helped to bridge the gap between older adults and the healthcare system and, through collective effort, ensured that their elderly parents were informed and involved in their healthcare decisions.

We also observed significant information-sharing within families. This was often facilitated through digital platforms like family WeChat groups (based on shared articles or videos), or occurred during family meals, where members discussed information they had gathered. This sharing process was not passive: it was an active strategy to ensure everyone was updated. Previous research has demonstrated that information sharing is common among family caregivers [30], who often disseminate the information they obtain to help others overcome barriers to information-seeking [31]. This dynamic is particularly evident when a patient or a family member faces language barriers in accessing essential information. Our research provides a clear example of this process, with an added layer of initiative in our context: older adults with language barriers often requested that younger family members search for local information on their behalf. In these cases, the younger generation acted as an assistant, while the older adults retained agency in information-seeking. In some other cases, when older adults obtained information independently through informal sources (e.g., friends or WeChat), adult children took on the role of verifiers, checking the credibility of what had been found. These differing scenarios illustrate how the level of agency among older adults shapes the nature of their children’s involvement—from supportive facilitators to active gatekeepers of health information.

Results reported here suggest that participant families generally preferred collective to individual healthcare decision-making. Two different strategies for shared decision-making were evident in people’s comments: either all members contributed to information-seeking and then reached a collective decision, or older parents described their symptoms, prompting adult children to seek information and bring it to the family for discussion and decision-making This finding is consistent with reported decision-making strategies in other behavioural domains; collaboration can include both joint activities and coordinated behaviour performed separately but in a synergistic manner [32].

Power dynamics were evident in the manner in which information was shared and used by participating families. Some adult children exerted control over their parents’ health, especially in managing chronic illnesses. Older parents recognised that this behaviour was consistent with caregiving rather than control. A study conducted in the U.S. [33] revealed similar findings; adults actively assisted their ill friends or family members to improve patient health outcomes. In this research, family members shared information in order to highlight what was “normal” or to raise concerns. Results suggested that both coercive and persuasive strategies were utilised to impact health decision-making. Expectations or “injunctive norms,” which highlight expected behaviour, were used to help guide the decision-making of the targeted family member [33]. We observed similar patterns in Chinese families in our study, although there were notable differences in parents’ reported reactions between the Chinese and U.S. contexts. In the U.S. study, some families reported clashes between family members over health information. The person with the health condition attempted to maintain control and to influence family members’ attitudes by challenging the credibility, accuracy, and relevance of shared information. Conversely, our parent participants were generally cooperative with their children’s suggestions. This difference may be due to the unique circumstances of our study’s participants—older adults with language barriers, limited access to local networks, and high dependence on their children in navigating a foreign health system. In contrast, the U.S. study involved participants who, according to the authors, had greater language proficiency and familiarity with the health system, which may have enabled more assertiveness or resistance in health-related discussions or have resulted in a more strong focus on individualism. These contextual differences likely influenced the observed variations in intergenerational power dynamics.

A recurring observation in our study was an implied desire, expressed by some adult children, for their parents to exercise greater autonomy in health decision-making. Although traditional Chinese family values emphasise collectivism and mutual responsibility, younger generations in our study seemed to encourage a shift towards more independent healthcare engagement for older adults. For example, F5 adult child (see Section 4.1) noted that they hoped their parents could make more informed decisions independently without over-relying on family support. This reflects a subtle generational shift in decision-making, where the younger generation advocates for their parents’ autonomy alongside interdependence. Interestingly, some parents also expressed a wish for autonomy in seeking health information and making decisions independently. However, this desire was constrained by the practical barriers, such as language limitations and unfamiliarity with the Australian healthcare system, which made independent engagement challenging. Suggestions for empowering parents to make independent decisions are discussed in Section 5.3.

The observation that information is linked to power in families is important. Offspring in our study acted as gatekeepers, ensuring that their parents had access to appropriate information. This reflects Foucault’s theory of power and control over information, [34] where expertise and surveillance allow one group to exert influence over another. Our study provides vivid examples that highlight how, in the context of immigrant Chinese families, these power dynamics shape how health decisions are made and whose voices are prioritised.

Intergenerational conflicts were described in the study despite the perceived benevolence of the children’s participation. Chinese families resolved internal conflicts by turning to healthcare providers, who were trusted across generations. This reliance on healthcare professionals helped mitigate potential disagreements within the family because their opinions were perceived as authoritative. However, while our findings suggest strong trust in health professionals among these families, existing research shows that trust is highly context-dependent and varies within and across countries and cultures (e.g., [35–40]). Trust in healthcare providers can fluctuate based on clarity of communication, accessibility, prior experiences, and institutional credibility —not merely cultural background [37, 39]. We encourage future research to examine how interpersonal and systemic trust develops in migrant families navigating unfamiliar health systems, rather than assuming uniform patterns across cultures.

#### Variability in practice across Families

Although collective decision-making was commonly described in our interviews, there was notable variability in who ultimately made the final decision. These differences reflected two main dynamics: a more distributive approach, where decision-making was collaborative, with all members contributing equally; and a more traditional hierarchical structure where older parents delegated decision-making to their adult children.

Some families adopted a more distributive decision-making process, with input valued from all members and a strong emphasis on autonomy for the person whose health was impacted. Family 13 exemplified this, with a guiding principle that “whoever’s body it is, they make their own decisions, even if it’s a child”. This type of family valued participation without overstepping boundaries, maintaining a clear distinction between individual autonomy and collective decision-making.

On the other hand, in families following the parent delegation with consent model, older parents typically delegated decision-making authority to their adult children, who took on the role of guiding and suggesting health decisions. However, parents still retained the authority to consent to, or veto, decisions, as seen in Family 8. This dynamic mirrors a blend of traditional values and the practical realities of living in a new cultural context. In this type of family, paternalism still plays a significant role, with the patriarch or another senior family member representing the family authority. Traditionally, this role was filled by the male head of the household (Wang, 2021). However, in modern scenarios such as in our study, we also saw female figures, such as the daughter in Family 12, who was a nurse. She was regarded as an authority by other family members, and took on the role of primary decision-maker due to her medical knowledge, thus altering traditional power structures.

This shifted power dynamics based on knowledge highlights a key point: family decision-making is not static. Instead, it is fluid, shaped by the evolving needs of the family and the availability of knowledge. This highlights how decision-making is influenced not only by cultural values [41] but also by practical necessities, ultimately shaping how families navigate the Australian healthcare system.

### 4.2 Interactions of Chinese families with the Australian Health system

Approaches to healthcare in Australia often assume that patients act as autonomous individuals, for example, in reacting screening invitations or participating in preventive behaviours. However, for many Chinese older adults, health decisions are negotiated collectively within the family and health information must also be available in their preferred language to be effective. When such family decision-making and information-seeking dynamics are overlooked, individualised approaches may fail to engage older adults effectively.

Chinese families in Australia often face challenges when adapting to the local healthcare system, primarily due to the differences between Australian and Chinese healthcare models. In Australia, patients must first consult a General Practitioner (GP) to determine the need for specialist care, sometimes leading to lengthy waiting times, even for people with private health insurance. In contrast, healthcare in China is more immediate, with patients able to access specialist medical attention directly. Additionally, those interviewed parents with personal connections at local hospitals indicated greater confidence in their health decision-making. By contrast, those families with health insurance in China highlighted discussions about returning to China for treatment.

These health system differences often left older parents feeling unfamiliar and confused about the Australian healthcare system. Unfamiliarity extended to crucial areas such as insurance management and medication procurement. As a result, adult children played a pivotal role in guiding their parents through the complexities of the Australian healthcare system. As F11 adult child mentioned, “besides medication, they rely on me for most medical decisions because they don’t fully understand the local healthcare system.” This reliance underscores the gap in understanding and highlights the need for interventions like community seminars to help older adults gain greater independence in managing their healthcare while living in Australia.

In addition to family support, friends and community networks significantly shaped healthcare decision-making for Chinese families. Friends’ recommendations were often the first source of information, providing a trusted and effortless way to access validated advice based on personal experiences. This led to the adoption of similar healthcare practices within these networks, such as choosing the same General Practitioners (GPs), the same insurance plans, opting for the same vaccinations, or participating in health screenings like bowel cancer checks after hearing positive feedback from others who had undergone the procedure. This finding resonates with recent studies [42] that have shown that interventions involving peer/community health workers and regular counselling are particularly effective in CALD populations, including Chinese families.

Digital platforms, particularly WeChat group chats, also function as online community hubs, serving as vital channels in facilitating information-sharing within these communities. These group chats, typically consisting of Chinese residents living in the same region in Australia or speaking the same dialect, are useful for disseminating timely updates on available vaccinations, upcoming health events, and other relevant community activities. Further utilisation of online information sharing could serve to foster a sense of solidarity among community members and ensure that families remain informed and make healthcare decisions collectively. The use of these digital platforms further exemplifies the dynamic of collaborative and collective decision-making, where both family members and the broader community actively engage in navigating the healthcare system together.

### 4.3 Implications for Practice

Researchers have emphasised the need for more nuanced approaches to provide accessible information that caters to the preferences of patients, such as source and presentation style, rather than simply focusing on content [43]. In this section, we provide examples and suggestions tailored to the specific context of Chinese families living in Australia.

#### Enhancing language support for immigrant populations

Language barriers pose a significant challenge for Chinese immigrants in Australia, affecting both older adults and younger generations, who may use English in their daily work, but struggle with medical terminology. This challenge makes accessing healthcare effortful and can lead to delays or misunderstandings. To alleviate these issues, healthcare providers should offer services in multiple languages. This could involve not only providing multilingual staff, translated materials, and interpreter services, which are already available through initiatives such as the Translating and Interpreting Service (TIS National). But also, these resources must be accessible, trusted, and widely used. Several participants in our study noted that government websites and authorised in-language materials exist, but they are often difficult to locate, overly formal, or perceived as lacking authenticity. Improving dissemination through community-based channels, as well as tailoring the style and quality of translations, is therefore crucial to make these resources more effective.

Participants in our study noted that although government websites and authorised sources often provide translated materials, these translations are frequently difficult to understand and may lack authenticity. Therefore, it is crucial to improve the quality of translated health information to make it more accessible and user-friendly for non-native English speakers.

#### Supporting older adults’ autonomy in health decision-making

Although family-based decision-making was described by many Chinese Australian families in our study, several older parents expressed a strong desire for greater autonomy in managing their own health (e.g., see the quote from F9 parent in Section 4.2.2). They articulated a wish to attend medical appointments independently, make their own decisions, and reduce their reliance on their children — particularly when adequate language support and system familiarity were available. This desire for autonomy did not reflect a rejection of collective values, but a nuanced negotiation of independence within an interdependent family structure. This highlights the importance of designing health systems that empower older immigrants to engage directly with care processes, when they choose to do so.

To support this autonomy, we encourage healthcare services to design services that enable older adults to make and attend appointments independently, offer training for staff on how to support older clients in navigating systems respectfully, and provide optional support tools—such as simplified navigation guides or bilingual health assistants—so that older adults can choose their preferred level of support.

By recognising and supporting the diverse roles older adults wish to play in their own care, healthcare systems can honour both cultural expectations and individual preferences—contributing to more inclusive and empowering care models [41].

#### Culturally tailored health education programs

Interviewed families highlighted the need for culturally-tailored health education programs (e.g., small group sessions) [44–47] to help the immigrant Chinese families, especially their older parents, to understand the local healthcare system and obtain health-related information. Chinese families’ preferences for, and reliance on, community networks, highlighted their potential for more extensive utilisation. Apart from providing online resources, regular seminars and workshops are appreciated because they bring people together and bridge the knowledge gap, offering clearer guidance on navigating the Australian healthcare system, understanding preventive care practices, and accessing available resources.

#### Leveraging WeChat for community-based health initiatives

As the dominant social media platform in the Chinese community, WeChat plays a crucial role in connecting individuals, sharing information, and facilitating daily communication. Given its importance, healthcare providers can leverage WeChat to disseminate accurate and timely health information. This can be done by publishing news, events, and health-related content through official WeChat accounts and WeChat video platforms - an article and video platform built in the WeChat application. Additionally, organising and managing group chats on WeChat to spread information within specific communities could enhance the reach and impact of health campaigns.

#### Addressing intergenerational health decision-making dynamics

The interdependence between older parents and their adult children in making health decisions highlights the need for healthcare providers to consider family dynamics in healthcare delivery [47]. Training healthcare professionals to engage both generations in discussions and respecting the collaborative decision-making processes could lead to more effective and satisfactory healthcare experiences. This approach also potentially solves problems with intergenerational conflict that can arise in discussions about health because our findings indicate that Chinese families value doctors’ opinions highly, often using them as a strategy to reach consensus on health matters. However, strict legal requirements mean that clinicians are generally obliged to communicate directly with the patient unless another adult holds legal authority (e.g., enduring power of attorney). This may restrict collective decision-making; easier access to such authorisations could help bridge this gap.

### 4.4 Limitations

This study, while providing valuable insights into the decision-making dynamics within Chinese immigrant families in Australia, does have some limitations. First, the study may not fully represent the diversity within the Chinese immigrant population. The sample size, although sufficient for exploratory purposes, does not include the full spectrum of socio-economic backgrounds, regions of origin, or varying degrees of integration into Australian society.

Another limitation relates to the evolving nature of family dynamics and decision-making processes. As younger generations become more integrated into Australian society, their influence within the family structure and their approach to decision-making may shift over time. Our study captures the current state of these dynamics, but it may not fully account for how these processes could change in the future. Specifically, we did not explore in depth the length of time younger generations have lived in Australia, which could be a significant factor in shaping their decision-making roles and integration levels. Future research could benefit from a more detailed examination of these variables to better understand the long-term evolution of family dynamics.

Additionally, the focus of this study was primarily on decision-making within families. Other important influences require further investigation, including the impact of peer networks, healthcare providers, and media. These external influences could play a critical role in shaping health decisions, particularly in immigrant communities, where trust in external sources can vary widely.

## 5. Conclusion

This study explored complex health decision-making processes within Chinese immigrant families in Australia, revealing a blend of collective approaches, and individual input, across different family dynamics. These approaches help family members, especially older parents, overcome challenges such as language barriers and unfamiliarity with the Australian healthcare system. Decision-making process is influenced by the roles that family members play, and the power dynamics are shaped by variations in the capacity to seek relevant information. Our findings underscore the need for culturally sensitive public health strategies that accommodate these diverse dynamics [41, 47].

## Data Availability

The interview transcripts generated and analysed during this study contain potentially identifying and sensitive personal information and cannot be shared publicly due to ethical and privacy restrictions. De-identified excerpts relevant to the findings are included within the manuscript and its Supporting Information files. Researchers who meet the criteria for access to confidential data can request access to the full de-identified dataset by contacting the Human Research Ethics Committee of the University of Melbourne (contact via) and the corresponding author.

## Notes

### Competing Interest Statement

The authors have declared no competing interest.

### Funding Statement

Yes

### Author Declarations

This research was approved by the Human Research Ethics Committee of the University of Melbourne (Ethics ID number: 26567).

